# “Endless Opportunities”: A qualitative exploration of facilitators and barriers to scale-up of two-way texting follow-up after voluntary medical male circumcision in Zimbabwe

**DOI:** 10.1101/2023.12.20.23300319

**Authors:** Chelsea Elkins, Sandra B. Kokera, Phiona Vumbugwa, Jacqueline Gavhera, Kathleen M. West, Katherine S. Wilson, Batsirai Makunike-Chikwinya, Lewis Masimba, Marianne Holec, Scott Barnhart, Sulemana Matinu, Beatrice Wassuna, Caryl Feldacker

## Abstract

In Zimbabwe, the ZAZIC consortium employs two-way, text-based (2wT) follow-up to strengthen post-operative care for voluntary medical male circumcision (VMMC). 2wT scaled nationally with evidence of client support and strengthened follow-up. However, 2wT uptake remains suboptimal. Understanding the gap between mobile health (mHealth) potential for innovation expansion and scale-up realization is critical for 2wT and other mHealth innovations. Therefore, we conducted an exploratory qualitative study with the objective of identifying 2wT program strengths, challenges, and suggestions for scale up as part of routine VMMC services. A total of 16 in-depth interviews (IDIs) with diverse 2wT stakeholders were conducted, including nurses, monitoring & evaluation teams, and technology partners – a combination of perspectives that provide new insights. We used both inductive and deductive coding for thematic analysis. Among 2wT drivers of expansion success, interviewees noted: 2wT care benefits for clients; effective hands-on 2wT training; ease of app use for providers; 2wT saved time and money; and 2wT strengthened client/provider interaction. For 2wT scale-up challenges, staff shortages; network infrastructure constraints; client costs; duplication of paper and electronic reporting; and complexity of digital tools integration. To improve 2wT robustness, respondents suggested: more staff training to offset turnover; making 2wT free for clients; using 2wT to replace paper VMMC reporting; integrating with routine VMMC reporting systems; and expanding 2wT to other health areas. High stakeholder participation in app design, implementation strengthening, and evaluation were appreciated. Several 2wT improvements stemmed from this study, including enrollment of multiple people on one number to account for phone sharing; 2wT inclusion of minors ages 15+; clients provided with $1 to offset SMS costs; and reduced SMS messages to clients. Continued 2wT mentoring for staff, harmonization of 2wT with Ministry e-health data systems, and increased awareness of 2wT’s client and provider benefits will help ensure successful 2wT scale-up.

## INTRODUCTION

Voluntary medical male circumcision (VMMC) is a critical component of comprehensive HIV prevention and scaled nationally across much of Sub-Saharan Africa.[1] VMMC is safe with few serious adverse events (AEs).[2–8] Post-operative follow-up mitigates the risk of AEs and ensures quality care. In accordance with global guidance, national VMMC guidelines recommend in-person follow-up visits in the two weeks following procedure.[9] However, with few AEs, in-person follow-up compounds pressure on already constrained systems in resource-scarce settings, like Zimbabwe, with severe national healthcare worker (HCW) shortages.[10]

The International Training and Education Center for Health (I-TECH) formed the ZAZIC Consortium (an acronym composed of letters from member partners: Zimbabwe Technical Assistance, Training and Education Center for Health (Zim-TTECH); Zimbabwe Association of Church Related Hospitals (ZACH); and Zimbabwe Community Health Intervention Research Project (ZiCHIRe)) to conduct VMMC in partnership with the Ministry of Health and Child Care (MoHCC).[10] ZAZIC has been a leader in VMMC innovations since 2013, working to integrate VMMC services into routine healthcare delivery and ensuring quality service delivery by exploring and addressing factors that contribute to AEs.[2,11,12] ZAZIC’s VMMC program is efficient and safe, reaching over 500,000 males with an AE rate of 0.13%.[6]

In response to few AEs and required post-operative visits that burdened overstretched VMMC providers, I-TECH designed, tested, evaluated and scaled an app using two-way texting (2wT) to improve the quality of post-operative care while reducing the burden on HCWs and reducing costs. The 2wT system was rigorously tested via a randomized control trial (RCT) in 2019, providing evidence on 2wT safety, usability, and workload reduction.[10,13–15] In brief, the 2wT system for VMMC is a mobile health (mHealth) innovation that was built using the open-source Community Health Toolkit (CHT). This 2wT system is optimized for local Zimbabwean users, allowing VMMC clients to opt into post-operative follow-up via SMS (text message) with VMMC nurses instead of scheduled, in-person, post-operative visits.[16] 2wT provides automated educational messages about the healing process and facilitates interactive SMS with a VMMC nurse for concerns or reassurance, triaging clients to in-person care when needed. 2wT supports VMMC scale-up by allowing the vast majority of interviewees to heal at home while those in need of in-person review for potential AEs are promptly referred to on-site care by the 2wT nurse. Since 2019, while overcoming the restrictions and complications of service delivery within the COVID-19 context, ZAZIC transitioned the 2wT follow-up approach from the RCT to national scale-up, reaching over 33,000 men by 2023.[17] 2wT was also successfully evaluated in a RCT in both rural and urban South Africa, with similar findings in safety, usability, efficiency, and improved quality of follow-up.[18–20]

2wT remains a safe and effective follow-up method, with high usability among providers and clients: over 50% of all ZAZIC VMMC clients receive 2wT-based follow-up. 2wT adheres to the mHealth principles [21] and aligns with recommendations gleaned from recent mHealth literature [22–25], including: 1) 2wT follow-up is brief, reducing risks of poor retention, including challenges related to phone theft/loss; 2) 2wT’s hybrid approach of one-way educational messages to meet client desires for action-oriented knowledge combined with interactive, client/nurse telehealth improves engagement in care; 3) VMMC context reduces confidentiality concerns; 4) locally-led, human-centered design (HCD) tailors content to meet client and HCW needs; 5) SMS meets expressed client preferences while WhatsApp-like interface for providers reduces concerns of low digital literacy; 6) and, the “Hub-and-Spoke” model provides a dedicated central 2wT nurse, known as the “Hub Nurse”, to support site-based teams (“spokes”), reducing response delays.

Despite building upon digital health successes, support for scale-up, and clear evidence of 2wT advantages, the pace of 2wT expansion remains lower than expected. 2wT is not intended to meet the needs of all males in all VMMC locations. Yet, adequate network coverage and high phone ownership suggest that uptake should be closer to 75% of all VMMC clients. Elucidation of some of the causes and possible strategies to help 2wT fulfill its promise could identify barriers and potential solutions for other evidence-based apps aimed at improving client care while reducing HCW workload – a critical issue across many low- and middle-income countries (LMIC).

Qualitative research that engages diverse stakeholders, including HCWs, can help better understand app dissemination opportunities and challenges.[24] Soliciting widespread user feedback as apps move from research to routine practice is also in line with World Health Organization (WHO) guidance to promote mHealth equity and provide quality evidence for decisions surrounding expansion, scale-up, and institutionalization.[22,26,27] In the mHealth field, especially during COVID-19, many apps did not transition from research or niche implementation to routine practice.[28] Moreover, users who are given multiple opportunities to provide feedback may be more invested and engaged in digital health programs, an important consideration as 2wT continues to generate multi-level buy-in for expansion.[14,19,29,30] User feedback is not a goal that can be accomplished, but rather a core component of frequent and flexible adjustment and adaptation to specific programmatic context.[19] Encouraging insights and feedback from HCWs, in particular, raises the likelihood of understanding what works in real-world clinical settings, aiding implementation success.[31] As 2wT has successfully been brought to scale and maintained for over 2 years in the Zimbabwe setting, there is a unique opportunity for inquiry to share with others in the growing mHealth field.

To improve the current program as well as inform future 2wT scale-up efforts, we conducted an exploratory qualitative study among 2wT stakeholders, including 2wT supervisors, enrollment nurses at 2wT VMMC sites, ZAZIC collaborators, and staff from 2wT technology partner, Medic Mobile. The main objective of the study was to identify 2wT program strengths, challenges, and suggestions for 2wT scale up as part of routine ZAZIC VMMC services. Engaging this broad group of digital health users is critical to inform successful scale up from RCT to widespread use in the routine VMMC setting. Incorporation of more voices from LMIC public health settings also enriches the optimization process and further improves fit of digital health where these innovations are most needed.

## METHODS

### Setting

The ZAZIC Consortium provides comprehensive VMMC services in partnership with MoHCC in 14 districts, with a mix of urban, peri-urban and rural sites in Zimbabwe. VMMC demand creation and service delivery are also aimed at populations at higher risk of HIV, like artisanal mining and high migratory communities. We interviewed 16 2wT program partners for this study. We defined a *2wT program partner* as an individual from an organization that participated in the design, implementation, scale up, and/or maintenance of ZAZIC’s 2wT program for VMMC follow-up.

Interviewees included staff in a variety of roles across the ZAZIC Consortium; employees from the program’s technology partner, Medic Mobile; and MoHCC nurses implementing 2wT on site. ZAZIC and MoHCC site nurses are all based in Zimbabwe’s Harare, Midlands, Mashonaland Central, Mashonaland East, Mashonaland West, Masvingo, Matabeleland North and South Provinces. Medic Mobile employees interviewed are located in Kenya, Ghana, Uganda, and Nigeria.

### 2wT Intervention at scale

The 2wT system has been described in detail previously.[14,15,17] Within the ZAZIC program, clients are enrolled into 2wT on the day of VMMC by VMMC site teams. On the day of VMMC, 2wT clients receive augmented post-operative counseling about how to interact with the 2wT system and reminders on signs of complication to monitor during the follow-up period. After the procedure, clients receive a daily text with an educational message or a prompt for a response. On interactive prompt days, clients have the option to respond with “0” (no issues) or “1” (possible problem), indicating whether they have concerns about their healing. A 2wT nurse interacts with all males who report a problem or those who write and SMS into the 2wT system with a concern. If the men do not respond with a concern, there is no immediate follow-up. Men who do not respond at all by day 7 are contacted with a call to confirm there are no issues.

Clients can send an SMS message to the 2wT system at any time. After-hours coverage on weekends and holidays ensures a timely response for urgent matters, with the regular 2wT nurse responding during routine clinic hours. Figure 1 (below) illustrates the 2wT interaction flow.[10] The 2wT implementation operates as a Hub (the central 2wT “Hub Nurse”) and spokes (all sites), referred to as a “Hub-and-Spoke model”. Sites enroll 2wT clients and conduct follow-up visits, completing forms on in-person visits and observed AEs. The 2wT Hub Nurse interacts with men from all sites on a daily basis, providing reassurance, triaging men to in-person reviews using a “referral for care” form, and contacting men who do not respond by SMS or phone. The Hub Nurse refers men who have not responded to sites for follow up on Day 8.

**Figure 1:**
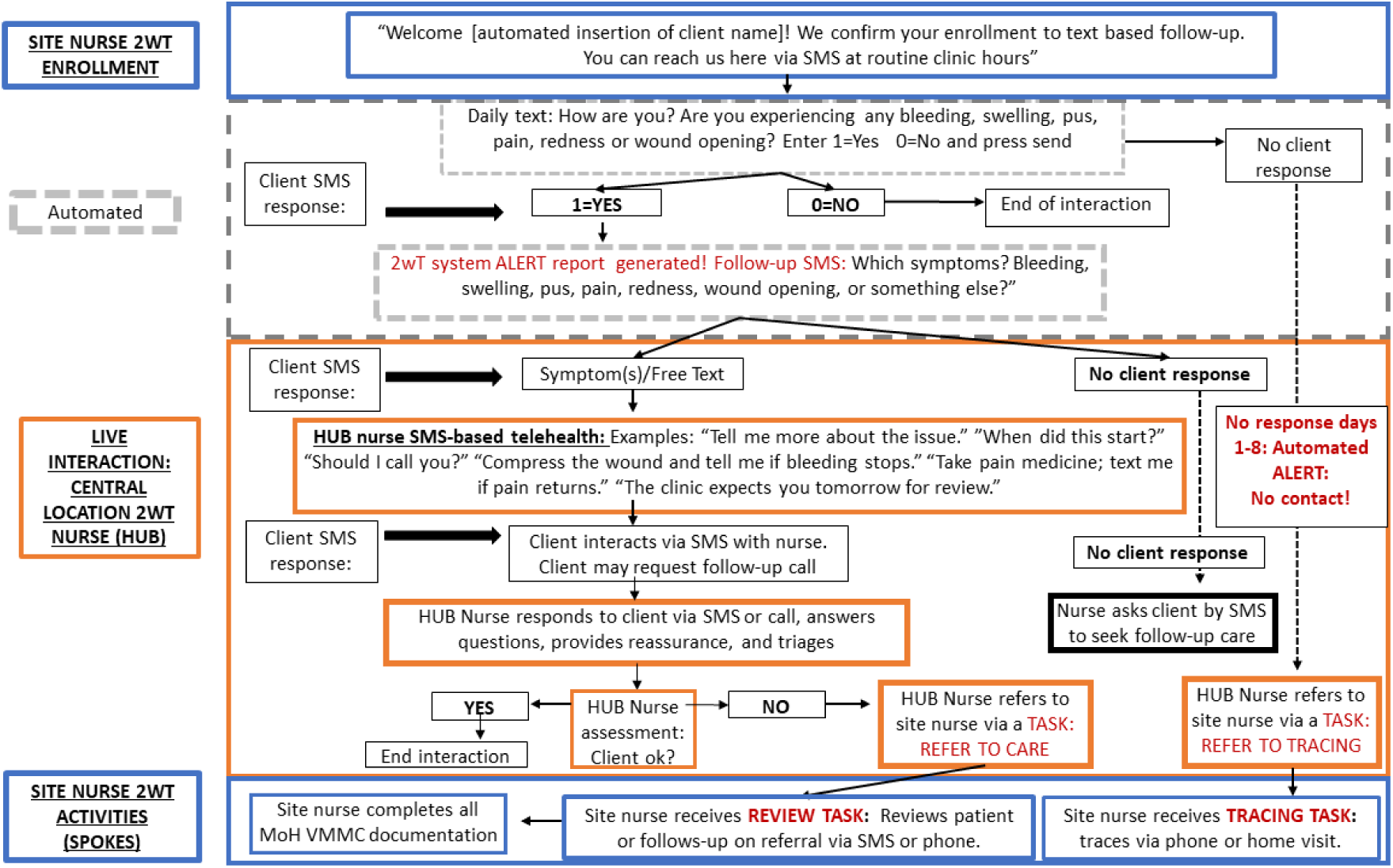
2wT System VMMC Follow-Up Flow[10].

### 2wT Training for scale-up

For scale-up, 2wT training was conducted by a core team of four ZAZIC HCWs over a 1-week period and included 30 administrative staff from the MoHCC national-, province-, and district-levels. The training was cascaded down to facility levels by ZAZIC teams with two nurses per site attending a two-day training. Trainings were conducted virtually or in-person on-site, depending on COVID-19 protocols and logistics issues. In both formats, training included facility nurses, data clerks, site managers, and demand creation staff. The ‘trainer-of-trainers’ approach with on-the-job mentorship was used: the first group of trained staff subsequently trained other staff at their sites as staff turned over. The 2wT training curriculum includes theory and practical sessions conducting post-operative counseling; enrolling clients; documenting visits and client tracing outcomes into the 2wT system; and routine monitoring and evaluation. In-person trainings resumed immediately after COVID-19 precautions were reduced.

### Data Collection

We conducted interviews from December 17^th^, 2021 to February 27, 2022. We selected 16 interviewees, using a combination of purposive and snowball sampling from ZACH, ZICHIRE, I-TECH, MoHCC and Medic Mobile teams who provided direct management, support, or implementation of 2wT at national-, district-, or health facility-level.[10,32] The sample size is reflective of research demonstrating that a high quantity of interviews is often not necessary for high caliber qualitative research,[33] as well as a recent review of qualitative studies that noted studies with 9-17 individual interviews can be sufficient for thematic saturation.[34] Furthermore, our goal was to hear from key implementing partners in the 2wT program, and we successfully interviewed the majority of partners (16/21) across all organizations that supported the program implementation. We conducted and recorded the individual in-depth interviews (IDIs) via Zoom or WhatsApp using a semi-structured interview guide.[21] Interviews were conducted remotely due to COVID-19 health concerns. Most interviews lasted 20-35 minutes, with an average length of 27 minutes. The lead researcher transcribed the interviews, editing for clarity as needed.

### Data Analysis

We undertook a thematic analysis approach.[35] We iteratively developed a codebook, initially identifying codes deductively based on study goals and interests. Additional inductive, or open, coding allowed us to capture unanticipated ideas regarding program barriers.[36] After several iterations, a program member reviewed and commented on the draft before we finalized the codebook. The primary coder (CE) applied the codebook to all transcripts using ATLAS.ti version 9.[35,36] A secondary coder (SK) reviewed the primary codes and identified additional open codes and minor discrepancies in interpretation that were resolved through a common writing process.[37] The study team identified, a priori, three domains according to study goals. We sorted codes into these domains and examined the coded data for patterns within and across stakeholder groups, to develop into themes under each domain. Again, we undertook a member checking process with individuals in the program, as well as the larger study team, to ensure accurate interpretation and communication of ideas.

### Positionality and trustworthiness

The lead interviewer is not employed by Zim-TTECH, nor any partner organization, but is affiliated with the University of Washington Department of Global Health, a funder of the 2wT project. This may have impacted how interviewees responded to questions. The remaining authors are either from, or work closely with, the Zim-TTECH organization which may have influenced their interpretation of the responses. All researchers were attentive to iterative process evaluations of trustworthiness,[25] assessing and strengthening the study’s methodological rigor, transparency, and inclusion across data collection, analysis, and results reporting phases.

### Ethics

The human subjects division of the University of Washington’s Internal Review Board determined that these human subjects research activities are for the purposes of quality improvement that qualifies for exempt status (IRB ID: STUDY00014561). All program partners provided verbal consent to be interviewed and recorded prior to participating in this study.

## RESULTS

### Interviewee Characteristics

Among 21 partners contacted, 16 participated in the study, representing the five organizations responsible for the 2wT implementation process. Interviewees either worked for the technology partner, Medic Mobile (n=5); as MoHCC site nurses (n=2); or within the ZAZIC Consortium (n=9), including Zim-TTECH (n=3); ZACH (n=3), and ZiCHIRe (n=3). Most interviewees (n=11) lived in Zimbabwe, except for Medic Mobile interviewees who were based in Kenya, Ghana, Nigeria, or Uganda (Table 1).

**Table 1:**
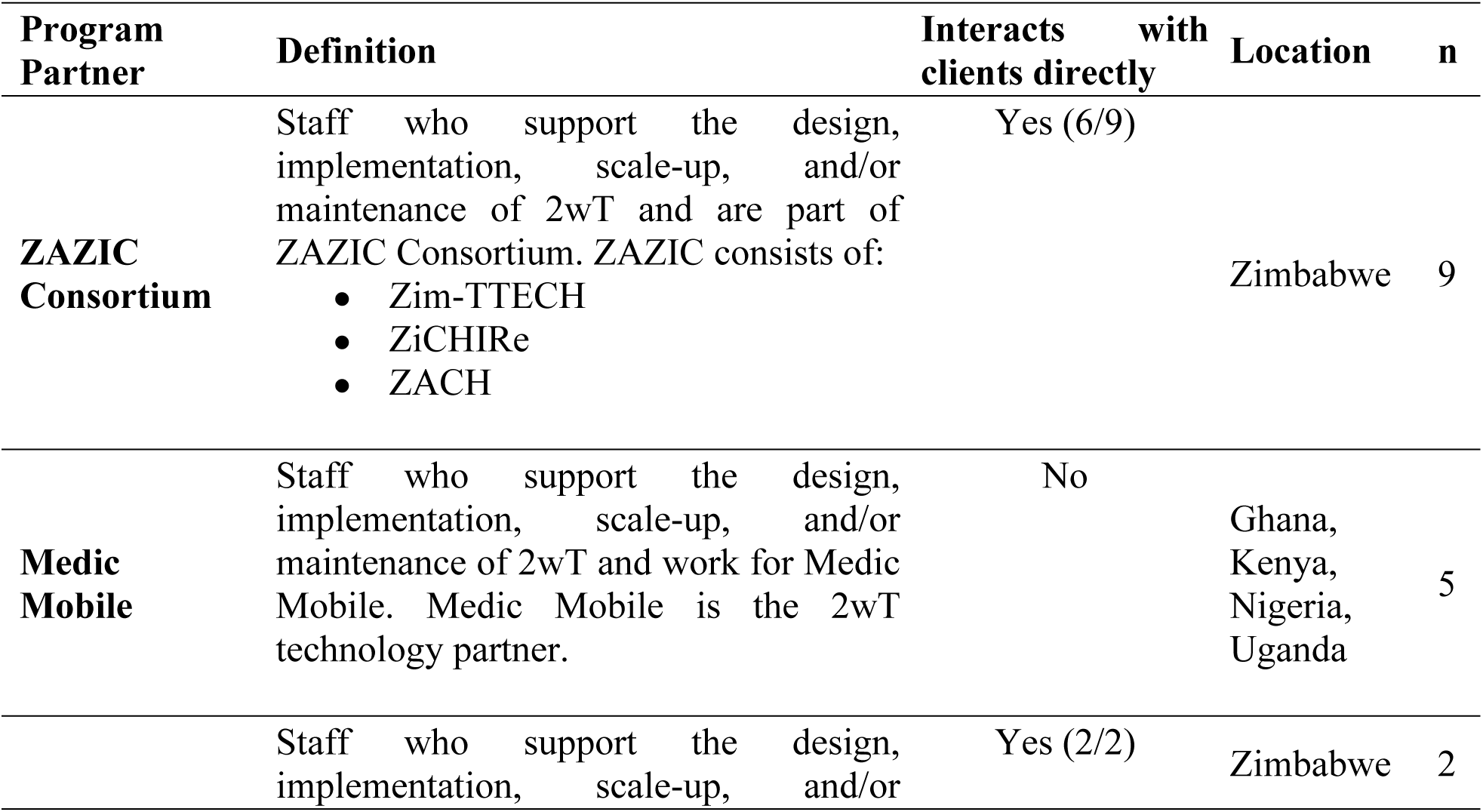
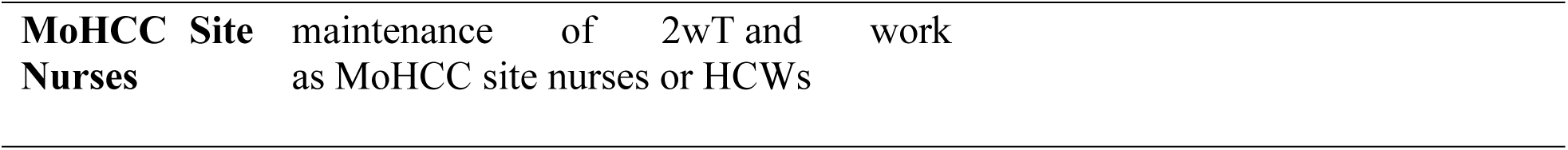
Characteristics of Participating Program Partners (N=16)

| Program Partner | Definition | Interacts with clients directly | Location | n |
| --- | --- | --- | --- | --- |
| **ZAZIC Consortium** | Staff who support the design, implementation, scale-up, and/or maintenance of 2wT and are part of ZAZIC Consortium. ZAZIC consists of:<br>• Zim-TTECH<br>• ZiCHIRe<br>• ZACH | Yes (6/9) | Zimbabwe | 9 |
| **Medic Mobile** | Staff who support the design, implementation, scale-up, and/or maintenance of 2wT and work for Medic Mobile. Medic Mobile is the 2wT technology partner. | No | Ghana, Kenya, Nigeria, Uganda | 5 |
|  | Staff who support the design, implementation, scale-up, and/or | Yes (2/2) | Zimbabwe | 2 |

MoHCC site nurses conduct 2wT related tasks, such as client recruitment, reviewing clients referred for suspected AEs or loss to follow-up, and 2wT oversight at the health facility-level. ZAZIC partner responsibilities include facilitating the 2wT training; providing overall 2wT system operations oversight, serving as the 2wT “Hub” to triage clients for care, following up on clients lost to follow-up or with suspected AEs, referrals for in-person follow-up, monitoring 2wT system challenges, maintaining documentation, and conducting quality assurance. Technology partners at Medic Mobile provide technical support to the 2wT program, including development of software, upgrading 2wT systems, and supporting continued program staff and HCW use. Our results are organized by three domains: Program Strengths, Program Challenges, and Suggested Improvements, with themes under each domain as presented in Table 2 (below).

**Table 2:** Results Domains and Corresponding Themes.

| Domain 1: 2wT Program Strengths | Domain 2: 2wT Program Challenges | Domain 3: Suggested 2wT Improvements |
| --- | --- | --- |
| Sub-themes<br>• 2wT improves quality of client follow-up care<br>• Continuous 2wT training + mentoring is effective<br>• 2wT saves costs for providers, program and clients, creating enthusiasm for scale<br>• The 2wT app is easy for HCWs and clients to use<br>• 2wT is widely accepted by clients, communities and MoHCC | Sub-themes<br>• Lack of 2wT trained or motivated HCWs at site level<br>• Persistent structural technological barriers<br>• Client struggle with SMS cost and SMS frequency<br>• Complexity of data management between MoHCC paper forms and 2wT digital database | Sub-themes<br>• Technology improvements for HCWs and clients<br>• Train more 2wT staff<br>• Replace paper data reporting with 2wT digital M&E<br>• Increase MoHCC ownership and transfer to local e-health systems<br>• Expand 2wT to other health programs |

### Domain 1: 2wT Program Strengths at Scale

### 2wT improves quality of client follow-up care

Interviewees overwhelmingly reported that the 2wT program is largely a success, as one interviewee remarked, “*we have a lot to really celebrate.*” [IDI-15] HCW interviewees noted advantages for client care, since now *“[clients] have the motivation and are willing to respond…People are interested in their own health and are willing to communicate directly with the two-way texting nurse.”* [IDI-01] HCWs expressed that 2wT increases close post-operative monitoring and improves identification of AEs over in-person visits:

> *“[2wT] has increased the follow-up rate. It also has improved adverse event surveillance. I think that close interaction with clients would always [allow us] to know if there is an adverse event.” [IDI-07]*

2wT also appears to improve follow-up data quality since the system both stores interactive messages and alerts nurses for timely documentation of referrals to care outcomes. Instead of clerks transcribing clinical notes from multiple sources, the 2wT nurse completes reporting. “*With two-way texting…it’s the service provider who is in contact with the client. [They are] getting first-hand information and entering the data.” [IDI-09]*

#### Continuous, on-the-job 2wT training is effective for 2wT scale-up support

Interviewees described the 2wT curriculum content of in-class role plays, 2wT-based practical exercises, and on-the-job training, as simple and useful. “*The training has been quite successful in giving all the information within a day, and [HCWs are] able to use it soon afterward*.” [IDI-04] The one-day format caused little disruption to the clinicians’ duties, and the practical training enhanced learning the content. One interviewee noted, “*hands-on [training] sped up the learning process, and also improved the buy-in and appreciation of the technology.*” [IDI-01] When COVID-19 restrictions led to a temporary change to virtual training, HCWs missed on-site support and repetitive practice. Still, virtual training did have upsides, because “*all players are there, everyone is there, they exchange ideas. [Training by] visiting HCWs in their facilities takes time and people don’t have one platform to exchange ideas.”* [IDI-08] Interviewees remarked on the importance of continuous trainings, and voiced that the role of the Hub Nurse should “*[provide] continuous mentorship support”* [IDI-08]

#### 2wT saves provider, client, and program costs, creating enthusiasm for continued scale-up

Interviewees reported that the 2wT program contributed to resource savings, both financial and human. For HCWs, 2wT saved travel time over in-person follow-up, leaving time for demand creation or other health services. On the program level, 2wT also decreases costs, “*because for you to go for follow-ups, you need a vehicle, you need to support the service providers…it involves a lot of personnel. But if there’s someone just sitting there and in charge of the clients [via 2wT], it’s a lot easier.”* [IDI-02] Respondents noted that 2wT largely eliminates the need for clients to travel and take time off work. “*Two-way texting came at a time when clients, especially adult males, were not willing to have the commitments to meet the service delivery teams and have them conduct reviews. So, I think it’s popular. It’s very convenient for clients.*” [IDI-07] Several HCWs confirmed that client demand for 2wT has grown, so that “*clients will just come in and they’ll ask, ‘Oh, I want to be part of the clients that will be texting back and forth. I won’t be able to come [into the clinic] because I’ll be at work.”* [IDI-03]

#### The 2wT app is easy to use, making it attractive for healthcare workers

For HCWs, most interviewees identified the ease and usability of the 2wT app as a major strength of the program: *“It’s just very simple steps to follow.…. It’s actually very friendly for the healthcare staff.”* [IDI-03] HCWs also remarked on the multi-lingual 2wT system that reaches clients in their preferred language and that it is easy for clients to use, regardless of their education. “*The [clients] easily understand it, even those who we were not sure would be able to use it because of literacy levels.”* [IDI-06]. The simplicity of the app for HCW and client users was attributed to the 2wT co-design approach. A respondent from technology partner, Medic Mobile, noted: “*We designed a system with our users, for our users…. That’s something that I would say I’m really proud of, just from the start: making sure that the digital health intervention really resonates with its users and that it’s really solving for their pain points.”* [IDI-15]. Interviewees also appreciated ongoing 2wT adaptation and revision based on ongoing user feedback. For example, to improve messaging speed, Medic Mobile developers leveraged the Community Health Toolkit (CHT) integration with another open-source tool, RapidPro, “*to allow more [2wT] messages to be sent out at a given time. I think that’s probably one of the big things…that might be going well with the scale-up.”* [IDI-16]. Frontline HCW appreciated this consistent effort to simplify and improve their own user experience, that of their VMMC clients.

#### 2wT is widely accepted by clients, communities and MoHCC

Most interviewees reported that the 2wT program was generally well received by clients and HCWs alike, resulting in a high program uptake. “*The response rate has been good. Even in areas where there is no good [cellular] network.*” [IDI-01] Interviewees identified promotion of the 2wT follow-up method at community and facility levels as a program strength, including distribution of 2wT branded masks and hats, sensitization on 2wT during community health talks, 2wT training for village health workers, and VMMC outreach to adult working men highlighting availability of 2wT-based follow-up. As a result, one interviewee stated that “*two-way texting is now a household name.*” However, HCWs noted that broad support was not immediate. HCWs initially had their own doubts about client safety and concerns about clients’ digital literacy, but with time, “*our population kind of embraced [2wT].”* [IDI-01] While success among clients and HCWs was encouraging, respondents were most impressed with MoHCC acceptance. “*The fact that the Ministry really appreciated this two-way texting intervention and supported the scale up [is a] huge success*.” [IDI-15] Another respondent considered MoHCC partnership as critical for success, noting that “*[2wT is] a good innovation [which] is telling policymakers that these [physical] follow-ups are not necessary, and policy should be informed by this finding.”* [IDI-02]

### Domain 2: 2wT Approach Challenges for Expansion Maintenance

#### Difficult maintaining core numbers of 2wT trained or motivated HCWs at site level

Multiple respondents reported lack of sufficient trained HCW to support the 2wT program. Although sites had at least one trained staff member, those with only one found that enrolment was disrupted if the trained nurse was not on duty or took a different job. *“Once our trained nurses have gone away, we have no one enrolling our clients.”* [IDI-04] Multiple partners identified staff shortages due to attrition, or a shortage of trained HCWs at sites, as a constraint to scale-up. However, even with on-site 2wT training, staff must perceive 2wT as part of routine duties, and not as a special or additional task. Multiple interviewees pointed out that some sites had HCWs who appear reticent to use to 2wT since, “*in some sites, you would get someone who is not very keen with two-way texting*.” [IDI-02]

#### Persistent technological barriers to sustained 2wT roll-out

Poor cellular network connectivity and access to devices slow 2wT expansion. Of the 16 program partners interviewed, 13 discussed poor mobile networks as a constraint to scale-up that deprived program partners and clients access to the system. Especially in rural areas, *“some of the areas are very remote and network coverage is very poor…It’s difficult to implement when there is no mobile network.”* [IDI-07]. Still, urban areas could face power outages that affect network availability. Interviewees also reported that the 2wT app was slow when high volumes of new clients were added. “*After a while, the system starts to slow down because of the volume of data on it. [Now], we purge old data from the devices, so the system doesn’t have to filter through so much data before it loads.”* [IDI-14] System delays can lead HCWs to enroll clients after hours, causing syncing issues that result in data gaps, or, infrequently, some clients not receiving messages on time or at all.

#### Clients struggle with 2wT costs and communication

Interviewees identified client challenges, such as low technology literacy and phone sharing. Some clients did not have access to a phone or shared a phone with others in their household -- a particular challenge in rural areas or with the enrollment of minors. IDI-09 commented that, *“Most of them share phones with their parents, their siblings, and sometimes you may get responses, not from the client, but from another person.”* Multiple interviewees also stated that cost was a potential barrier as clients had to purchase their own airtime for communicating with the nurses via 2wT. *“The issue of just the availability of the dollar, to be texting back and forth [is a challenge].”* [IDI-03]

#### Complexity of data management between MoHCC paper forms and 2wT digital database

To date, MoHCC mandates paper reporting for VMMC procedures, resulting in data duplication for 2wT clients. IDI-02 remarked on *“the duplication of the system in the enrollment. Because we have paper-based client intake forms, and we have this two-way texting system, which is collecting the same data.”* IDI-03 identified the additional work this created, noting the need *“for the hard copy for the circumcision itself. As much as we are going to enter them into the system, we are still going to enter them in another physical form.”* Making changes to global or national policies such as discontinuation of paper forms is not simple. “*There is not much control in terms of determining the pace of moving things. It’s all reliant on the Ministry of Health.”* [IDI-13]

### Domain 3: Suggestions to facilitate wider scale-up of the 2wT approach

#### Improve the technology for HCWs and clients

Several interviewees stated that previous clients should be archived to free the system memory and increase the application speed on HCW devices. Features such as improved data visualization and audible notifications for new messages could promote timely nurse responses. Several interviewees proposed 2wT be delivered on WhatsApp, versus traditional SMS, “*because with WhatsApp, [clients] say it’s cheaper, and they no longer communicate using an SMS.* [IDI-05] Additionally, multiple interviewees discussed needing system updates to allow multiple clients to enroll with the same phone number or f multiple phone numbers per person, accommodating those with multiple SIM cards. *“The platform should allow a number to be entered twice, especially when the surname is the same, to show that these [clients] are related and they’re using the same number.”* [IDI-11] Lastly, considerations of improved security, such as two factor authentication for access to 2wT client data were voiced as being needed.

#### Train, and refresher train, more 2wT staff

While almost every interviewee stated that the 2wT training was an overall success, some cautioned, *“if you are to expand, there is a need to cascade the training down, so that you can also have provincial-based trainers, or district, or even site-based trainers.”* [IDI-07] Incorporating the 2wT program training into the standard VMMC training package for all new VMMC staff could also ensure that all MOH service providers become skilled with 2wT: *“Because when we, if we, are going into sustainability, that is going to be a huge barrier*.*”* [IDI-11] Lastly, improving the training’s reach by extending 2wT training to community health workers, and offering consistent refresher training, will help fill personnel gaps: “w*hen one [trained healthcare worker] is absent, or unwilling, another can take over.*” [IDI-06]

#### Replace paper with 2wT for all VMMC data

Duplication of paper and electronic data increases HCW burden, potentially compromising data completeness in either system. *“Maybe if we could do away with the paper based and we just go digital, it might improve our adverse event reporting?”* [IDI-04] Another interviewee suggested that if all VMMC data were entered into the same system, 2wT enrollments could be automated, and those not enrolled into 2wT could still have their routine VMMC data entered digitally. *“Those with phone numbers automatically go into two-way texting. Those without, you have to indicate that they didn’t bring a phone. Electronically moving away from the use of paper [client intake forms].”* [IDI-11] Comprehensive electronic VMMC data could ensure data linkage “*to other databases that are used by Ministry of Health as well, so that it’s available to a wider group,”* [IDI-08], making data more readily available to decision-makers.

#### Transition 2wT to more MoHCC ownership and integrate 2wT into Zimbabwean e-health

Several respondents noted a desire to offer 2wT to other VMMC implementing partners and transferring implementation to central MoHCC leadership. 2wT could be *“made a national thing, through the Ministry of Health through sensitization…to say, now everybody’s using two-way texting through the Ministry of Health, I think that would improve uptake.”* [IDI-04] The open-source (no fee) CHT 2wT software gave respondents optimism for potential MoHCC transition, “*because we were worried that if its proprietary software, once the other partners would be off [stop funding 2wT], the Ministry of Health would not be able to sustain its use.”* [IDI-13] To support transition, respondents noted that local software engineers from partner organizations and/or MoHCC must be trained to competence and confidence in the CHT, including the 2wT system, and that 2wT would need local server hosting. *“Transiting software engineering skills to the local team for 2wT maintenance is a priority for consistently meeting the needs of end users.* [IDI15]. Lastly, there is a huge leap between recognizing 2wT benefits and integrating 2wT into MoHCC e-health architecture. *“Trying to sell [2wT] to the other people, especially the health informatics department, they appreciate it, but they had huge expectations that two-way texting would seamlessly integrate with all aspects of the electronic health system.”* [IDI-01] The pathway forward is neither simple nor straightforward.

#### Expand 2wT follow-up to other health conditions beyond VMMC

All respondents noted that 2wT could benefit clients and HCWs beyond VMMC. “*There are so many opportunities of which we haven’t done much in terms of demand creation… [2wT] information dissemination, there is a huge gap and opportunity.”* [IDI-07] For example, 2wT supported VMMC service delivery during COVID-19 and could be leveraged for subsequent pandemic waves. “*Two-way texting has come at the right time, because we are able to review clients without physical reviews… during this outbreak of COVID.*” [IDI-05] As COVID-19 wanes, but client demand for improved communication with HCWs grows, interviewees identified areas for 2wT growth. *“For example, growth monitoring, HIV care and treatment, immunizations and so on. It will be very useful because it informs the health worker how the client is and will keep the client informed about when to make a certain visit or take drugs.”* [IDI-06] Another interviewee suggested that 2wT can provide *“follow-up for outpatients with chronic conditions, say the diabetics or hypertensive clients, they can measure their own blood pressure and give feedback via 2wT and avoid unnecessary movements going back to the hospital or clinic.”* [IDI-07]

## DISCUSSION

This paper shares perspectives, insights, and suggestions from a unique set of 2wT app stakeholders, including frontline HCWs and software developers, to increase understanding of the process, pitfalls, and pathways forward to bring 2wT to scale in routine VMMC settings in Zimbabwe. These lessons learned should be of great interest in South Africa where 2wT is scaling after evidence of safety, usability, and efficiency gains.[18–20] Interviewees noted that drivers of 2wT scale-up included enhanced promotion of client care and cost benefits, comprehensive and continuous HCW training, and ease of app use for HCWs and clients. In contrast, lack of available HCWs, technology challenges, client communication costs, and complexities in the transition to MoHCC ownership stymied progress. Interviewees suggested that national 2wT scale-up would benefit from an expanded training cascade, transitioning from paper to digital VMMC reporting, MoHCC ownership of 2wT, and using the 2wT model for other short-term client follow-up. Several lessons learned merit additional consideration.

First, the 2wT training and mentorship model appears to play a critical role in creating cadres of confident, competent, 2wT HCW users. Similar to an evaluation from Sierra Leone demonstrating the critical importance of both initial and refresher training for HCWs for institutionalization,[38] 2wT respondents believed that the combination of brief (<1 day) on-site training with continuous and accessible mentoring was effective to engage providers and facility teams in 2wT implementation and maintenance. During COVID-19, remote technology-based training (Zoom or WhatsApp) was an advantageous stop-gap and fit easily into HCWs personal schedules.[39] However, it is clear that HCWs preferred on-site in-person training and follow-up mentoring when possible. To facilitate in-person training, more decentralized 2wT training cascades across more districts are needed to more swiftly respond to shortages of 2wT-trained HCWs and sustain implementation at scale.[40]

Second, as with other mHealth apps, 2wT enhanced provider to client communication and built relationships. As men typically underutilize healthcare services,[41,42] mHealth appears to strengthen the relationships between male clients and their HCWs.[17,19] HCWs are aware that men may not be proactive seekers of healthcare services and want to better engage their male clients in care.[43] 2wT helped clinicians better care for their rural and distant clients by providing a platform for timely client/provider communication,[44] engaging clients and providers in a shared responsibility for healing.[45] Our findings largely complement a recent review that found mHealth could improve coordination among HCWs, promote flexibility in follow-up care, and enhance relationships between clients and their care givers.[45]

While technology still presents some barriers, client and provider creativity can help overcome challenges reaching clients in more remote or less connected areas. 2wT is offline-first, allowing clients to enroll even without network coverage. Data are stored and then synced later when providers return to connectivity. Although 2wT requires that clients receive an enrollment message to confirm their correct phone number for follow-up, many facilities now utilize solar power and providers access facility WIFI to connect daily, minimizing delays in messages being sent and received. Provider confidence in reaching clients is maintained despite short-term delays in messaging. Moreover, even in areas with poor connectivity, clients are aware of where to receive a signal near their homes or workplaces. Clients seek out these network “spot zones” for other communication needs and can receive and respond to 2wT messages concurrently, meeting the program reporting requirements and preventing client loss-to-follow-up.

Results from this qualitative study led to several program improvements since study conclusion. First, ZAZIC now enrolls minors aged 15-17 years in 2wT via the minor’s or the parent/guardian’s phone, expanding access to 2wT. Second, in response to client and provider feedback, daily text message with required response volume was reduced from 15 to 11 and language offerings were expanded to include Ndebele. These changes increased the educational content, reduced SMS volume overall, and responded to client communication preferences. Third, clients are offered $1 for initial transport reimbursement and to offset 2wT airtime costs, decreasing cost barriers to 2wT uptake. Fourth, system improvements allow clients to register with a primary and secondary number, responding from either number as network connectivity fluctuates. 2wT enrolls surgical and device-based VMMC clients (e.g., ShangRing), allowing men to have 2wT follow-up regardless of VMMC method. Lastly, technology improvements, such as weekly data purging and procuring phones with larger memory, have sped up processing for providers and facilities.

This study has several limitations. First, client perspectives from the scale-up phase were not included, but are warranted in future study. Second, interviewees who volunteered for this study may have had more time, capacity, and interest in the program than their counterparts who declined to participate. Additionally, while all partner organizations were represented, it is likely that views within partner organizations vary, and we likely did not capture the full range of views. Further study focusing on the technology perspective could further illuminate specific technical areas that were not well explored. Finally, these interviews took place over Zoom, sometimes with a poor connection, which may have led to some lack of clarity in transcription or reduced conversation comfort, potentially impacting the interview quality. Although these biases cannot be quantified, the research team feels that the strengths of this exploratory study outweigh the weaknesses, contributing understanding to mHealth scale up in Sub-Saharan Africa.

## Conclusion

There is rigorous evidence of 2wT’s positive impact on client care quality and acceptability among HCWs, but widespread scale-up remains incomplete. Despite enrollments of over 33,000 males ages 15 and older, and broad approval from diverse client, provider, partner, and MoHCC stakeholders, digital innovations still present complex challenges to further scale up. For long-term success, how and when to integrate 2wT into current e-health platforms, including potential integration into the Electronic Health Records (EHR) system, should be further explored. Continuous collaboration between mHealth program implementers across all VMMC partners, from ZAZIC to MoHCC, would help foster continuous 2wT improvements for clients and providers across districts. Although the 2wT app is free to use, highly skilled and paid workers are needed to adapt, maintain, and manage the system. In the short-term, ZAZIC is growing its capacity to independently manage the CHT-based 2wT app with support from Medic Mobile, creating a transition plan for local 2wT management, supervision, and maintenance on the pathway to MoHCC ownership. However, for longer-term 2wT transition to MoHCC, human resources, funding for SMS, server space, and availability of trained technology teams are required. If leveraged well, as noted by one respondent, 2wT could be utilized in a variety of settings and health conditions: “The opportunities are really endless.”

## Data Availability

Complete transcripts contain data that is sensitive or includes identifying information. The edited transcripts will be available on a case by case basis after reviewing all materials for any potentially identifying details. Interested researchers may contact the corresponding author, CF, at for copies of the transcripts. Or, researchers could also contact Jane Edelson, Regulatory Specialist at the UW, for access to the transcripts. Data described in the manuscript, code book, and analytic code will be made available upon request and completion of a relevant data-sharing agreement.

## Acknowledgements

The authors would like to thank the members of the ZAZIC Consortium, including the International Training and Education Center for Health (I-TECH,) the Zimbabwe Association of Church Related Hospitals (ZACH), Zimbabwe Community Health Intervention Research (ZiCHIRe), and Zimbabwe Technical Assistance Training & Education Center for Health (Zim-TTECH) for their support of 2wT. The authors would also like to appreciate the collaboration of the MoHCC and the facility teams for their dedication to quality VMMC service delivery, including 2wT based follow-up.

1. **Principles for Digital Development** [https://digitalprinciples.org/]

## Notes

### Competing Interest Statement

The authors have declared no competing interest.

### Funding Statement

The author(s) received no specific funding for this work.

